# Mapping suitability for Buruli ulcer at fine spatial scales across Africa: a modelling study

**DOI:** 10.1101/2020.04.20.20072348

**Authors:** Hope Simpson, Earnest Njih Tabah, Richard Phillips, Michael Frimpong, Issaka Maman, Joseph Timothy, Paul Saunderson, Rachel L Pullan, Jorge Cano

## Abstract

**Background:** Buruli ulcer (BU) is a disabling and stigmatising neglected tropical disease (NTD). Its distribution and burden are unknown because of underdiagnosis and underreporting. It is caused by *Mycobacterium ulcerans*, an environmental pathogen whose environmental niche and transmission routes are not fully understood. Active BU case searches can limit morbidity by identifying cases and linking them to treatment, but these are mostly restricted to well-known endemic areas. A better understanding of environmental suitability for environmental reservoirs of *M. ulcerans* and BU disease would advance understanding of the disease’s ecology and burden, and could inform targeted surveillance.

**Methodology/Principal Findings:** We used previously compiled point-level datasets of BU and *M. ulcerans* occurrence, evidence for BU occurrence within national and sub-national areas, and diverse environmental datasets. We fitted relationships between BU and *M. ulcerans* occurrence and environmental predictors by applying regression and machine learning based algorithms, combined in an ensemble model to characterise the optimal ecological niche for the disease and bacterium across Africa at a resolution of 5km × 5km. Climate and atmospheric variables were the strongest predictors of both distributions, while indicators of human disturbance including damming and deforestation, drove local variation in suitability. We identified patchy foci of suitability throughout West and Central Africa, including areas with no previous evidence of the disease. Predicted suitability for *M. ulcerans* was wider but overlapping with that of BU. The estimated population living in areas predicted suitable for the bacterium and disease was 29.1 million.

**Conclusions/Significance:** These maps could be used to inform burden estimations and case searches which would generate a more complete understanding of the spatial distribution of BU in Africa, and may guide control programmes to identify cases beyond the well-known endemic areas.

**Author summary:** Like many neglected tropical diseases primarily affecting the rural poor, Buruli ulcer (BU) is under-detected and under-reported within routine health information systems. As such, the burden and distribution are not fully known, impeding appropriate targeting of health resources, control, and care for people affected. Having previously evaluated and mapped the existing evidence for BU and its causative agent *M. ulcerans*, we concluded that the disease was likely to occur beyond the range of known endemic areas. However, we were left with the question of where exactly these undetected cases might be occurring. Answering this question required a more fine-scale approach: BU is highly focal, presumably due to local variation in the environmental factors which determine suitability for *M. ulcerans* survival and transmission to humans. We used the compiled evidence and geographical datasets to build statistical models representing the relationship between environmental factors and previously reported cases. This allowed us to define the ecological niche of BU, and subsequently to identify areas across Africa where this niche was met, providing suitable conditions for the disease. We constructed separate models of suitability for *M. ulcerans*, using locations where its DNA had been detected in environmental sources. Unsurprisingly, suitability for *M. ulcerans* was predicted to be wider than, but geographically overlapping with that for BU. This implies that beyond the conditions necessary for survival of the bacterium, additional factors are required for transmission to humans. The high-resolution suitability maps we present are intended to guide case search activities which may identify endemic areas beyond the known endemic range. Data on the true prevalence of BU from targeted case searches within predicted-suitable areas will also allow us to validate and refine the models, and potentially predict the actual probability of cases occurring within predicted suitable areas.

## Introduction

Buruli ulcer (BU) is a chronic necrotizing disease of the skin and soft tissue, which causes debilitating symptoms and sequelae, associated with a high burden of morbidity and stigma for patients and economic costs for affected households [1-3]. These impacts are felt particularly strongly in impoverished rural communities with poor access to health services [3, 4]. The infectious agent is *Mycobacterium ulcerans*, a slow-growing environmental bacterium which may be transmitted from aquatic environments to humans via inhalation or ingestion, or by penetration of the skin [1, 5, 6]. The main control strategy is active case finding in endemic areas to promote early case detection and effective treatment, which limits disease progression [7, 8]. BU occurs mostly in tropical and subtropical areas of West and Central Africa, with smaller foci in parts of Asia, South America, the Western Pacific and Australasia [9]. However, the disease is recognised to be underdiagnosed and under-reported, and may occur undetected in other parts of the world [9-12].

The distribution of BU is presumably linked to environmental suitability for *M. ulcerans* survival and replication, as well as to human and environmental factors favouring transmission [13]. On a continental scale, BU appears to be limited by climatic factors: it is restricted to tropical and subtropical regions and absent from arid areas [14]. Within endemic areas, the disease shows a highly focal distribution [15-17], but reasons for this are not well understood, since the precise niche and transmission routes of *M. ulcerans* have been difficult to characterise [18]. The pathogen has only been cultured from environmental and animal samples a handful of times [19-21], although it has been detected by PCR in aquatic environments of endemic and non-endemic areas, and a in wide range of potential hosts including mammals, fish, amphibians, and aquatic and terrestrial insects [22-26]. Consistent with the ecology of an environmental pathogen, the distribution of *M. ulcerans* in the environment appears to be wider than that of BU, suggesting that factors beyond environmental suitability for *M. ulcerans* are required for transmission [13, 14, 27].

Our understanding of the pathways of BU infection is also limited, partly by its long and variable incubation period, which makes it difficult for patients and clinicians to attribute particular events or activities to disease acquisition [28]. Local spatial analysis has identified several environmental variables associated with increased BU incidence, primarily proximity to rivers, as well as environmental disturbance and land-use changes including deforestation, urbanisation, agriculturalization and mining [17, 29]. Case control studies have identified contact with unprotected waterbodies as a risk factor for disease [30], suggesting that activities which bring people into contact with water sources harbouring *M. ulcerans* increase the risk of disease acquisition [31-33].

Given the recognised scale of BU under-detection and under-reporting, it is likely that BU occurs beyond the known range of reported cases. A better understanding of potential suitability for the pathogen in the environment and the disease in humans would help to improve its surveillance and control in countries where is known to be endemic. Furthermore, characterisation of the environmental factors linked to suitability for *M. ulcerans* and BU may reveal areas at risk for disease emergence, or places harbouring unrecognised cases.

In this investigation, we aim to identify environmental factors which characterise the environmental niche of *M. ulcerans* and BU disease in humans, and to model their respective relationships with BU. These analyses will be used to identify areas of continental Africa which may be suitable for *M. ulcerans* or BU based on their environmental characteristics.

## Methods

### Data on Buruli ulcer and *M. ulcerans* distribution

We used previously compiled spatial datasets of point locations of recorded occurrences of BU disease in humans, and of detection of *M. ulcerans* genetic material in biotic and abiotic environmental samples [9, 34]. Data for the final models was extracted from the database on 03/01/2020.

BU occurrence locations were restricted to those where BU infection was confirmed by a positive result for PCR targeting IS2404, or histopathology consistent with BU disease. To explore the model’s sensitivity to the case definition, we repeated the analysis using all locations where clinically diagnosed BU had been reported. We hereon refer to the two datasets as ‘*confirmed occurrences*’ and ‘*all occurrences*’ respectively.

The environmental dataset was restricted to locations where *M. ulcerans* DNA had been identified and distinguished from that of other mycobacteria: either by multiplex qPCR assays quantifying the relative copy numbers of IS2404, IS2606 and the KR-B domain [35]; byvariable nucleotide tandem repeat (VNTR); or mycobacterial interspersed repetitive unit (MIRU) typing [36, 37]. We hereon refer to this dataset as ‘*environmental occurrences*’.

All records were restricted to locations with reliable geographical coordinates and deduplicated by geographical location. Human and environmental locations were weighted by the year and the specificity of confirmatory tests reported (S1 Text).

### Environmental datasets used in ecological modelling

We assembled gridded datasets of 51 environmental variables considered relevant to the ecological niche of *M. ulcerans* [18]. All variables and their sources are shown in Table 1 and full details are provided in S3 Text.

### Variable selection

We compiled the gridded predictor variables at a resolution of 5km × 5km within a rectangular area of West Africa from latitude -13.57195, longitude-4.11032, to lat. 16.67107, long. 14.493. This area contained 94% of all BU occurrence locations, 95% of confirmed BU occurrence locations, and all environmental occurrence locations. We extracted the values of predictor variables at the locations of BU cases (all occurrences) and environmental occurrences of *M. ulcerans* DNA.

We used principal components analysis (PCA) to identify the minimum set of variables that best characterized the environment at observations of BU and of *M. ulcerans* from the global environment. We undertook separate PCAs on the human (confirmed cases) and environmental datasets, selecting variables that contributed most strongly to the minimum number of principal components collectively accounting for at least 80% of the total dataset variance.

### Pseudoabsence and Background data

The occurrence data used in the modelling framework were supplemented with systematically generated pseudo-absence and background data. The use of artificial absence data is a common approach in species distribution modelling, designed to account for geographically biased presence data and sparse absence data [38]. The terms background and pseudoabsence are often used interchangeably to describe artificial absence data, though Elith and Hijmans distinguish them on the basis that background data are intended to characterise the ‘environmental domain’ of the study-helping to account for geographical bias, while pseudoabsence data represent areas assumed to be unsuitable for the species and are intended to capture the environment in these areas [39]. We hereon use the terms in this sense.

Pseudoabsence and background points were both selected from a restricted geographical extent around occurrence points, defined by the spatial structure of the occurrence predictors. This has been recommended by previous authors as way to provide an ecologically meaningful definition of the study range [40]. Pseudoabsence points were sampled at higher density in areas of weaker evidence according to a systematic review of the geographical distribution of BU [41]. Background points were sampled at higher density around recorded occurrence points. More details on the generation of pseudoabsence and background points are provided in S2 Text.

Pseudoabsence and background weights were uniform within datasets and assigned so their sum was equal to the sum of occurrence weights in each model [42]. Human background points were restricted to a minimum distance of 10km from any occurrence location, and environmental background points were restricted to 10km from any human or environmental occurrence location.

The distribution of BU and *M. ulcerans* occurrences, pseudoabsence and background points are shown in S1 Figure and S2 Figure.

### Ensemble modelling

The environmental factors selected through PCA were used as predictor variables. We used the *biomod2* package in R [43, 44] to implement seven algorithms: generalized linear models (GLM), generalized additive models (GAM), generalized boosted regression models (GBM), artificial neural networks (ANN), multiple adaptive regression splines (MARS), maximum entropy (MaxEnt) and random forest (RF).

Individual model algorithms were each run 20 times with a random sample of 80% of data points, and evaluated with the remaining 20%. For each algorithm we calculated the mean true skill statistic (TSS), the mean positive correctly classified (PCC) and the mean area under the curve (AUC) of the receiver operation characteristic (ROC) [45]. Models with mean AUC above 0.8 were integrated in an ensemble using committee averaging to attribute higher weight to better performing models.

We plotted the importance values representing each variable’s contribution to the model and created marginal effect plots for the modelled covariates in the highest performing model ensemble.

### Estimating total population living in suitable areas

We calculated the total area suitable for BU, *M. ulcerans*, and the total area suitable for both, and extracted estimates of the population living in each of these areas from a raster representing estimated number of people per 1km^2^ grid square in 2020 [46].

## Results

### Datasets of BU occurrence in humans and *M. ulcerans* DNA detection in the environment

The modelled data included 2,183 unique point locations with reported cases of BU in Africa (Figure 1A). BU was confirmed by PCR or histopathology at 738 unique locations. There were 91 unique locations where *M. ulcerans* DNA had been detected by MIRU, VNTR or qPCR (Figure 1B). The dataset of clinically diagnosed human cases represented 16 countries, mostly in West and Central Africa, with a few in East and southeast Africa. The confirmed cases were restricted to 13 countries. The time period of human case detection was from 1957 to 2019. The median year of diagnosis was 2010. The 91 records of environmental detection of *M. ulcerans* represented three countries: Ghana, Cameroon and Benin, and covered the period from 2006 to 2018 with a median year of detection of 2013.

**Figure 1A:**
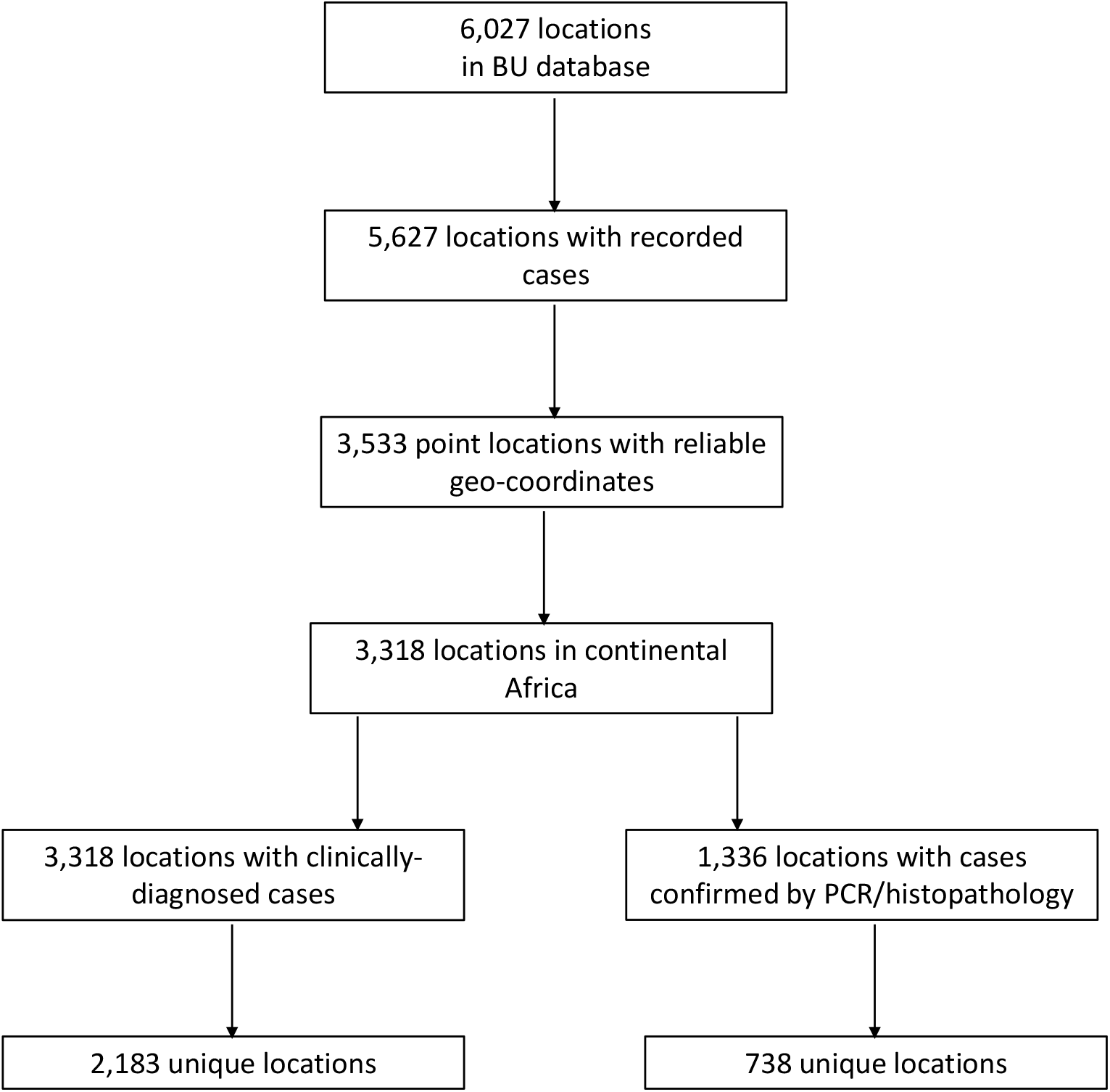
Selection of BU occurrence points from BU database.

**Figure 1B:**
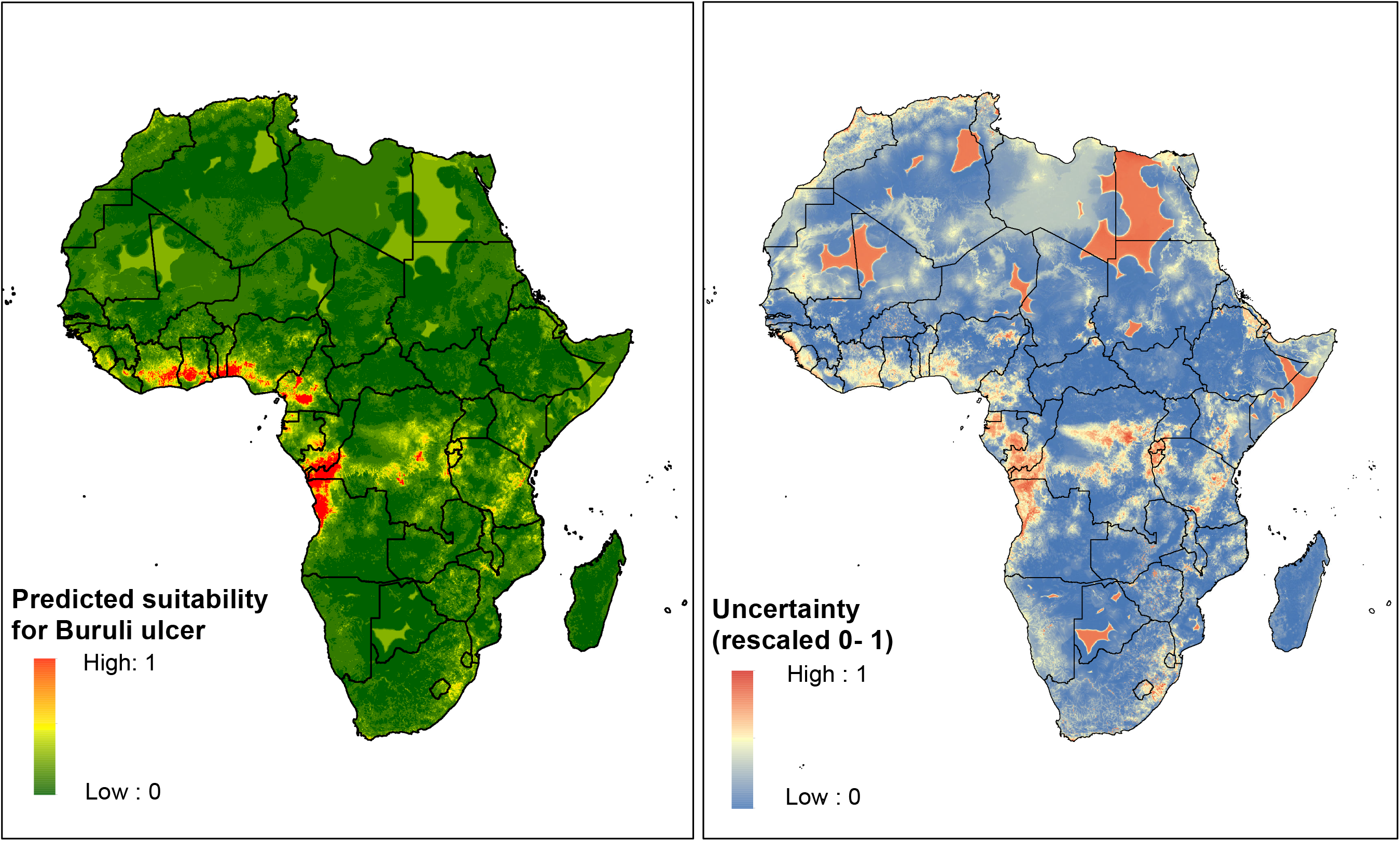
Selection of model environmental occurrence points from global database of MU occurrences.

### Principal components analysis

Ten principal components (PCs) collectively contributed 86% of variation in the human BU occurrence locations. Seven PCs characterised the environmental locations. The variables selected for each model are shown in S3 Text. The mean contribution of the most important predictors of human BU occurrence dropped sharply at 150km, while the contribution of the predictors of environmental detection of *M. ulcerans* declined at 300km (S4 Text).

### Environmental suitability for BU

All individual distribution models performed well with ROC scores above 0.8 (S5 Text). Mean PCC scores were between 79.3 and 91.6% and mean TSS scores were between 0.57 and 0.79 (Figures A and B, S5 Text).RF performed best with a mean PCC of 91.6%, a mean TSS of 0.79 and mean ROC 0.95. The final ensemble model showed an overall mean ROC of 0.96 with sensitivity of 87.1% and specificity of 92.9%. The mean TSS was 0.80 and the mean kappa score was 0.80 (Table 2).

**Table 2:** Validation metrics for ensemble models for BU and *M. ulcerans* suitability

|  |  | Weighted Mean | Lower CI | Upper CI |
| --- | --- | --- | --- | --- |
| BU suitability | TSS | 0.797 | 0.797 | 0.794 |
|  | ROC | 0.963 | 0.961 | 0.963 |
|  | kappa | 0.801 | 0.804 | 0.797 |
| MU suitability | TSS | 0.885 | 0.885 | 0.896 |
|  | ROC | 0.96 | 0.96 | 0.961 |
|  | kappa | 0.894 | 0.894 | 0.894 |

The minimum temperature of the coldest month was the strongest contributor to the RF models, followed by PET and distance to dams (Figure A in S6 Text).Environments with minimum temperature below 18°C were unsuitable, but suitability increased sharply at this temperature, decreased a little from 20-21°C, before increasing again, remaining high up to 24°C (Figure A in S7 Text). Optimal values of PET were between 1,000 and 1,500mm per month, corresponding to tropical rainforest canopy cover. There was a gradual decline in suitability for BU with increasing distance to the nearest dam up to a distance of 80km, after which suitability dropped rapidly.

The overall distribution was constrained to humid tropical areas and local scale variation appeared to be driven by hydrological features and deforestation patterns (Figure 2). The total area predicted to be suitable for BU was 338,500 km^2^, and the total population living in areas predicted suitable was 69.7 million (Table 3). Pockets of suitability for BU totalling >100 km^2^ were predicted in 16 countries in Africa, including all 12 countries along the west-central African coastline from Liberia to Angola (S2 Table). Angola had the widest area predicted suitable, followed by the Democratic Republic of the Congo, although the patches of suitability predicted in these countries were associated with high uncertainty. Nigeria had the largest population at risk, with 17.8 million predicted to be living in areas suitable for BU, followed by the Democratic Republic of the Congo where 10.8 million were predicted to be living in suitable areas (S2 Table).

**Table 3:** Total area predicted suitable and population in areas at risk for Buruli ulcer, M. ulcerans, and both, in Africa

|  | Total area<br>suitable (km<sup>2</sup>) | *Lower bound* | *Upper bound* | Population in<br>suitable areas | *Lower bound* | *Upper bound* |
| --- | --- | --- | --- | --- | --- | --- |
| BU | 338,500 | 283,950 | 425,600 | 69,740,630 | 62,573,091 | 83,050,754 |
| MU | 912,600 | 767,775 | 1,112,275 | 88,542,574 | 75,380,436 | 104,645,747 |
| BU & MU | 126,775 | 94,800 | 177,700 | 29,124,902 | 22,597,071 | 39,371,153 |

Suitability for BU and *M. ulcerans* is shown by country in S1 Supporting Maps.

**Figure 2:**
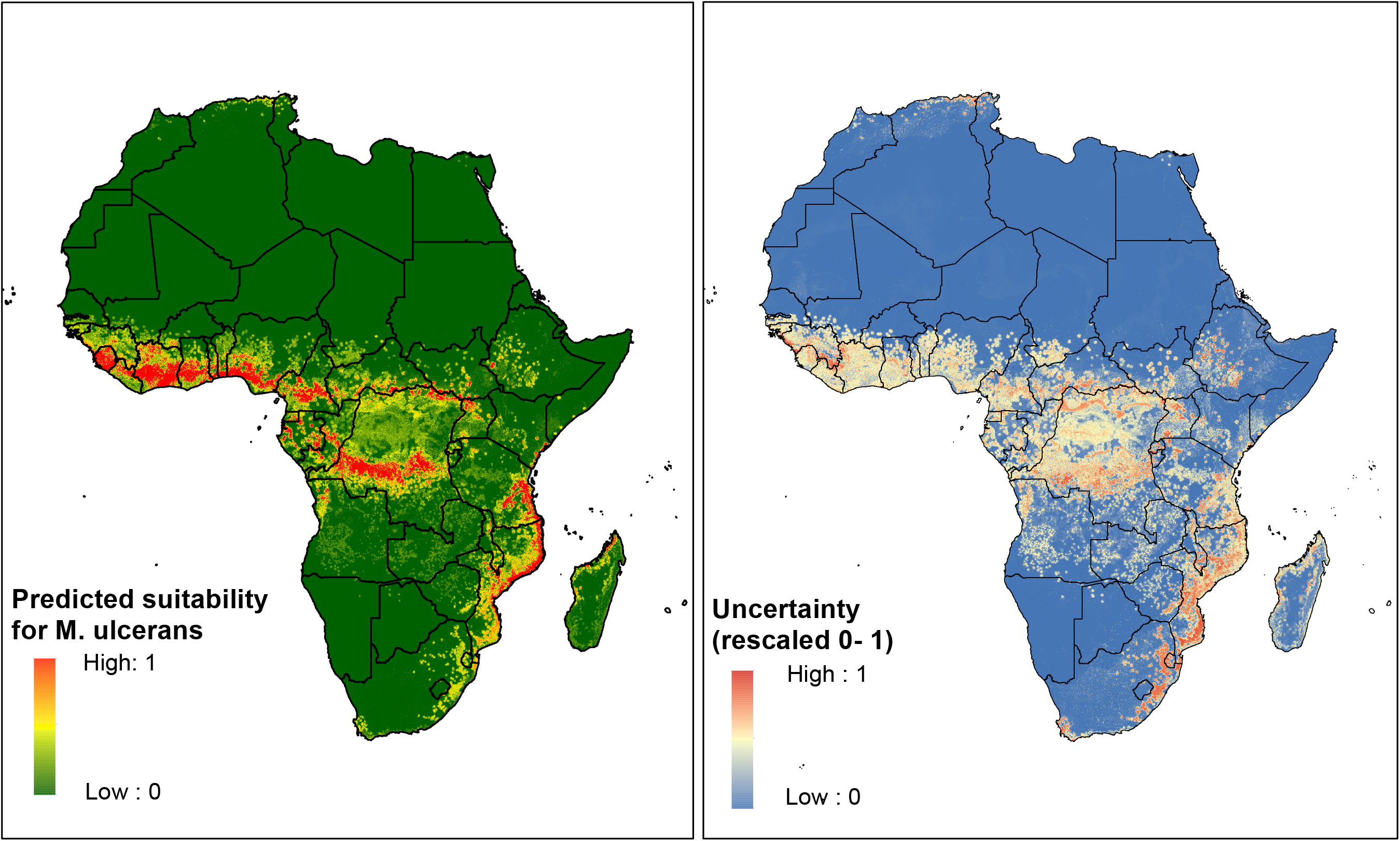
Predicted environmental suitability for the occurrence of BU disease in humans and associated error of prediction.

The model including all cases of BU (S3 Figure) gave similar results to the model including confirmed cases only. The Pearson coefficient of correlation between the two models was 0.86.

### Environmental suitability for M. ulcerans

All models performed well with ROC above 0.8, apart from MAXENT Phillips which was excluded from the ensemble model. Mean PCC varied from 0.80-0.89 between models and mean TSS was between 0.54 and 0.77 (Figures C and D, S5 Text). RF outperformed other algorithms in predicting the occurrence of *M. ulcerans*. The final ensemble model had a mean TSS score of 0.87, with a sensitivity of 92.3 and specificity of 94.5% (Table 2). The ROC sore was 0.99 and the kappa score was 0.87.

The minimum temperature of the coldest month was the strongest predictor of *M. ulcerans* occurrence in the RF models, accounting for 24.6% of all variance in the model (Figure B in S6 Text). Distance to deforested areas was also a strong predictor, accounting for 23.7% of the variance. Suitability was low at coldest month minimum temperatures below 18°C, and increased sharply to a peak at 25°C. There was a strong response to the distance to deforested areas, with high suitability at close range to deforested areas, decreasing sharply at 25km (Figure B in S6 Text).

The overall distribution appeared to be restricted by suitability for minimum temperature and precipitation in the driest month, with local variation driven by deforestation (Figure 3). The total area predicted to be suitable for *M. ulcerans* was 833,975km^2^, and the total population living in areas predicted suitable was 71.2 million (Table 3). Pockets of suitability were predicted in 31 countries (S2 Table). The DRC had the widest area predicted suitable (184,500 km^2^) followed by Côte d’Ivoire (120,600 km^2^).The highest population living in suitable areas was in Nigeria (26.7 million).

**Figure 3:**
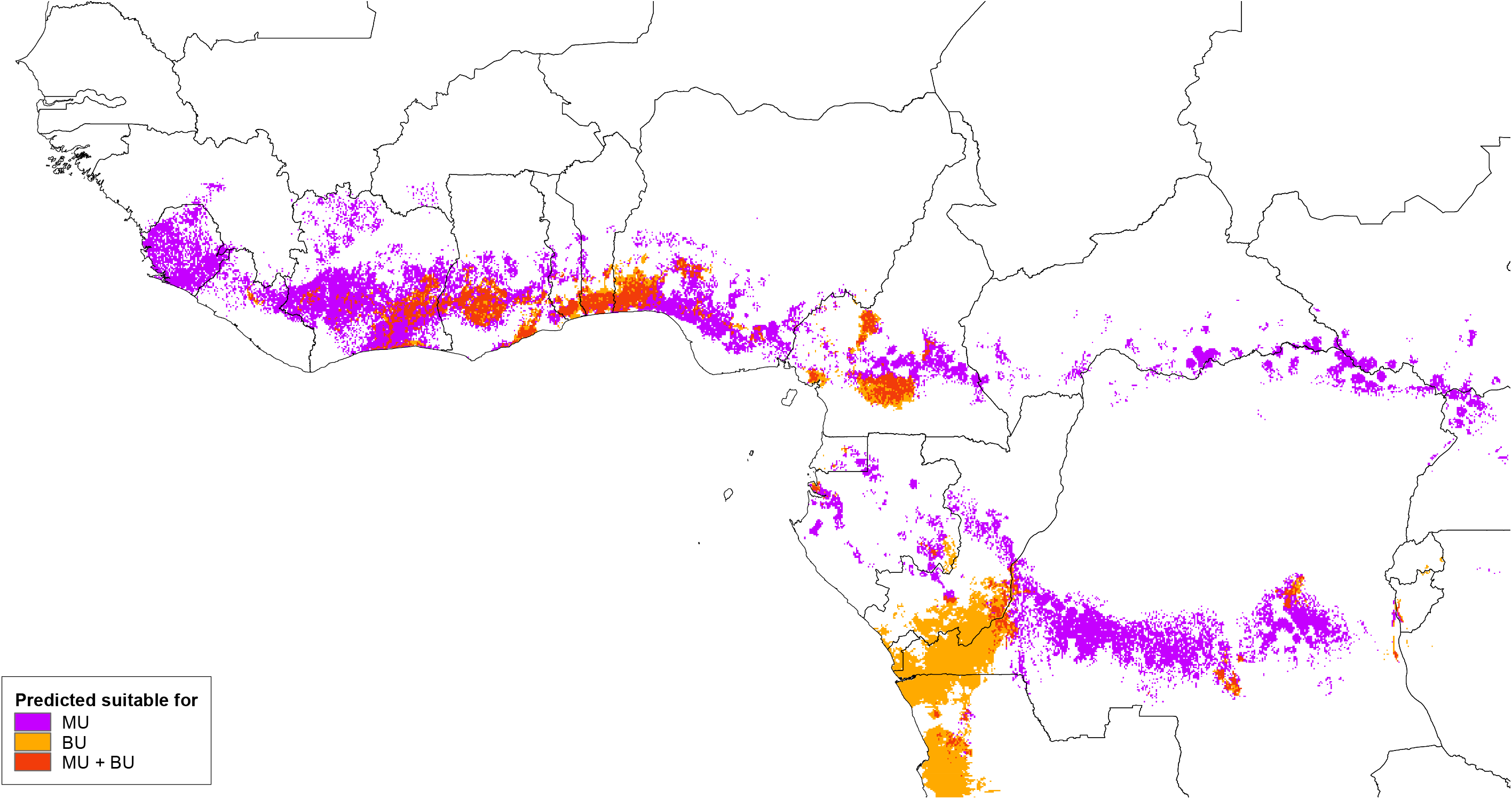
Predicted environmental suitability for the occurrence of *M. ulcerans* in the environment and associated error of prediction.

### Overlap of suitability for BU and *M. ulcerans*

The total area predicted to be suitable for both BU and *M. ulcerans* was 126,775 km^2^, with 29.1 million people predicted to be living in areas at risk. There were notable differences in the extents of the areas predicted suitable for BU disease and environmental *M. ulcerans* (Figure 4). There were wide areas in Sierra Leone and Cote d’Ivoire predicted suitable for *M. ulcerans* but not for BU disease, which was restricted to smaller pockets within these countries. Suitability for *M. ulcerans* was also predicted outside the predicted range of BU across central Africa and along the north-west cost of Mozambique, eastern Tanzania and coastal Kenya. In contrast, there were large patches in Central Africa predicted suitable for BU but not suitable for *M. ulcerans*. Ghana followed by Cameroon had the widest area predicted suitable for both BU and *M. ulcerans*. The highest populations living in areas predicted suitable for both BU and *M. ulcerans* were in Nigeria and Ghana, with 11.4 and 8.0 million respectively at risk.

**Figure 4:** Predicted overlap of environmental suitability for BU and of *M. ulcerans* occurrence.

## Discussion

We have used ecological niche modelling to identify environmental factors associated with the occurrence of Buruli ulcer and its causative agent *M. ulcerans*, and to predict environmental suitability for the disease and bacterium across continental Africa. Incorporating existing data on BU distribution and a geo-environmental definition of the range of occurrences, the resulting maps represent evidence-based predictions within a relevant spatial context.

There was substantial overlap in the factors contributing to suitability for human cases and environmental occurrence. Both BU and *M. ulcerans* were constrained to particular bioclimatic zones by environmental predictors which varied over large areas, characterising the humid tropical realm where BU is endemic in Africa. Our finding that areas with minimum temperature less than 18°C were unsuitable for BU supports evidence for a different epidemiology of the disease in Africa compared to endemic areas of temperate Australia and Japan [47]. Local-scale variation in factors including the distance to dams, deforestation and hydrology resulted in a patchy distribution of predicted suitability, consistent with our understanding of the epidemiology of BU, which is recognised to be highly focal in endemic settings [48].

We identified pockets of suitability for BU in patchy foci throughout the known-endemic range of the disease, particularly in the tropical zones of countries around the Gulf of Guinea. Throughout this range, suitability was predicted in regions not previously recognised as endemic. For example, two foci of suitability were predicted in Equatorial Guinea, which had no evidence of cases reported in peer-reviewed literature. However, these two areas correspond to the origins of a number of cases diagnosed by an expert in BU between 1995 and 2005 [49, 50].

Some locations in northern Cameroon with previous evidence of PCR confirmed BU were found to be unsuitable for the disease. This discordance may be due to the model’s failure to identify suitable environments in areas of lower BU incidence. However, given the great volume of surveillance data collected by the well-established BU control programme in Cameroon, some patients are likely to have been diagnosed outside the region where they acquired the disease [51], and we consider it plausible that some regions where BU has been recorded are not actually suitable for transmission. The suitability maps provide a depiction of areas potentially at risk for BU beyond what is known from the distribution of reported cases, currently the basis for targeting of surveillance and control. Given the recognised scale of underreporting of BU [41], the current approach is likely to exclude cases outside of known disease foci, and we suggest that areas predicted suitable for BU could be considered as targets for case finding activities, with the aim of identifying unrecognised foci and patients not known to the health system. Based on the wide areas of suitability predicted by this work and existing evidence of under-reporting of BU [52], the south of Nigeria would be a key target for case finding activities. There were wide areas predicted suitable in the Republic of Congo, the DRC and Angola, although these predictions were associated with significant uncertainty, which should be considered in the design of any future surveys. Suitable areas of Equatorial Guinea with historical evidence of cases would also be targets for case finding, although in this case the predicted suitable areas are more restricted, potentially necessitating a more stratified approach. A comparable approach has been applied to target malaria elimination efforts to transmission hotspots predicted through geospatial risk mapping [53], employing environmental modelling to impute risk in the absence of full surveillance coverage.

The model predictions could also be used to inform the design of cross-sectional surveys for BU, combining exhaustive case searches with environmental modelling to achieve robust estimates of prevalence. In a nationwide survey for podoconiosis in Cameroon, the selection of survey communities was stratified according predicted suitability for the disease based on a model trained mainly using data from Ethiopia [54]. This survey identified higher rates of podoconiosis in communities that were predicted suitable, implying a benefit in terms of the cost per case identified, compared to a survey employing random selection of survey communities.

The validation metrics we calculated demonstrate the ability of the models to predict BU and *M. ulcerans* occurrence with high accuracy. However, these measures do not indicate the models’ generalisability to areas beyond the range of known locations. Validation against external datasets would be required to assess this quality and is a target for future analysis.

The scale of analysis (grid cells at 5km × 5km) may have limited our ability to quantify the effect of predictors varying over small geographical scales. For example, the relatively small contribution of distance to waterways on suitability for BU was a surprising result, given that proximity to and contact with rivers have been identified as risk factors for BU disease [13, 18]. However, the variation we were able to capture in this predictor was limited, since most of the land area in tropical and sub-tropical Africa is within 10km of a river or stream, and much is within 5km. A previous analysis of land use and landcover and BU presence at large spatial scales in Benin found no association of BU at community level with proximity to water bodies [55]. The scale of analysis may also have limited our ability to capture fine scale variation in environmental suitability for BU. Our models predicted large contiguous areas of suitability in some areas with suitable bioclimatic conditions and within close proximity to stable night lights and deforested areas. Such areas may be suitable in reality, but exhibit an uneven distribution of disease due to factors not included in our models.

We have identified areas of high suitability for BU and *M. ulcerans* within known endemic-areas, and in areas not currently recognised as endemic, but with evidence of possible undiagnosed or misdiagnosed BU. The population at highest risk of BU is within areas where BU and *M. ulcerans* niches overlap, comprising almost 30 million people in 2020. The focal nature of BU distribution, the recognised scale of under-detection, and the impact of late diagnosis on disease severity strongly suggest a targeted approach to active case finding as a means to control this disease. The fine-scale, evidence-based predictions presented here could provide a tool to target such efforts, which will help to increase the proportion of cases linked to treatment, and contribute further research to establish the burden and distribution of this devastating disease.

## Data Availability

Data on Buruli ulcer and M. ulcerans distribution
We used previously compiled spatial datasets of point locations of recorded occurrences of BU disease in humans, and of detection of M. ulcerans genetic material in biotic and abiotic environmental samples.
These datasets are openly available.
Data for the final models was extracted from the database on 03/01/2020.

https://datacompass.lshtm.ac.uk/1143/

## Acknowledgements

The AIM Initiative was the sole funder of this work. We would like to recognise the contribution of all health workers, researchers and data managers who recorded cases of Buruli ulcer which were compiled into the global database of infections underpinning this study.

## Author contributions

RP and JC acquired the funding that enabled this project. JC conceptualised the study. JC and HS together developed the investigation and methodology-including the software (code for the analysis), and produced the visualisations presented. HS curated the data analysed, some of which was originally collected by other authors: ET, RP, MF, IM and JT. HS undertook the formal analysis and prepared the original draft. All authors critically reviewed the draft.

## Supporting Information

**S1 Text:** Weighting of occurrence points

**S2 Text:** Selection of background and pseudoabsence points S3 Text: Environmental variables used in modelling

**S1 Figure**: Distribution and weights of confirmed BU occurrence locations, background and pseudoabsence points

**S2 Figure:** Distribution and weights of environmental *M. ulcerans* DNA occurrence locations, background and pseudoabsence points

**S3 Figure:** Predicted environmental suitability for the occurrence of BU disease in humans and associated error of prediction, including all clinically diagnosed cases of BU

**S4 Text:** Spatial structure of occurrence predictors

**S5 Text:** Individual model performance evaluation statistics S6 Text: Variable importance plots for random forest models S7 Text: Marginal effect plots for random forest models

**S1 Table:** Total area predicted suitable and population living in suitable areas for Buruli ulcer, *M. ulcerans*, and both, by country in African continent.

